# Modeling the Effects of Routine Screening for Accidental Lab-Acquired Infections on the Risk of Potential Pandemic Pathogen Escape from High-Biosafety Research Facilities

**DOI:** 10.1101/2025.05.16.25327796

**Authors:** Byron Lev Mentis Cohen, Boyu Ren, William Hanage, Nicolas Menzies, Kevin Croke

## Abstract

Accidental lab-acquired infections (LAIs) risk releasing potential pandemic pathogens (PPPs) from BSL-3/4 facilities. We constructed a stochastic network infectious disease model to simulate how the probability of an outbreak of a pathogen resembling wild-type SARS-COV-2, following an initial LAI would be influenced by test-and-isolate interventions over a 100-day horizon. We varied test frequency (0–7 tests/week), peak sensitivity (50–100%), and isolation delay (0–3 days). For each of 192 parameter combinations, we conducted 1,000 simulations and used logistic regression to quantify how each parameter influenced the likelihood of an outbreak of 50 or more infections. Results indicated that even relatively infrequent routine testing significantly reduced the risk of outbreaks under diverse plausible scenarios, with greater reductions achieved at higher test frequencies. Once-weekly testing reduced outbreak risk by 52% under optimistic assumptions (80% sensitivity, 1-day delay) and by 29% under pessimistic assumptions (50% sensitivity, 2-day delay). Testing two and five times weekly yielded risk reductions of up to 62% and 71%, respectively, under optimistic assumptions, and 43% and 55%, respectively, under pessimistic assumptions. Logistic regression showed each additional weekly test decreased outbreak odds by 20%, each 10-point increase in test sensitivity reduced odds by 10%, and each additional isolation delay day increased odds by 15.5%. Interaction analyses revealed that longer isolation delays attenuated the protective effects of higher testing frequency and sensitivity. Routine lab-worker screening with prompt isolation substantially mitigates PPP escape risks. High-frequency testing has the greatest impact, and policymakers should consider implementing regular screening protocols.

## Introduction

Accidental lab-acquired infections (LAIs) with potential pandemic pathogens (PPPs) risk sparking a pandemic.^1–4^ Wet-lab research with PPPs is generally undertaken in Biosafety Level Four (BSL-4) or Biosafety Level Three (BSL-3) facilities, which operate at maximum and near-maximum biosafety levels, respectively.^5^ Such research can advance basic science and medical countermeasure development. ^6^ However, it also carries major risks of accidental human exposures and infections. From 1975 to 2016, there were more than 50 documented accidental human-caused exposures to high-consequence pathogens,^7^ including a smallpox outbreak associated with University of Birmingham’s lab in the UK in 1977, a 1977 influenza pandemic of presumed lab-related origin,^8^ anthrax infections near bioweapons facilities in the Soviet Union linked to more than 66 deaths in 1979, and four lab escapes of SARS-1 in Singapore, Taiwan, and China in 2003.^9^ Though the cumulative number of deaths from these events is modest, the mere existence of these events is an indicator that a biosafety lapse leading to escape of a PPP is plausible.

While the pandemic LAI risk posed by any particular research facility in a particular year is likely to be low, these risks are much higher when cumulated across time and over many research facilities. As of 2017, 14 laboratories were conducting research on highly-pathogenic avian influenza (HPAI) viruses modified for mammalian airborne transmissibility, notably H5N1 and H7N9, collectively termed by Klotz as the “matHPAI Research Enterprise.” Based on reported accidental pathogen releases from high-biosafety labs (2003-2017), Klotz estimated an annual release probability of 0.003 per lab, resulting in a cumulative five-year community release risk of 15.8% and pandemic risk of approximately 2.5%.^11^ There are reasons to believe the cumulative risk of escape of a PPP is likely higher than Klotz’s estimate, which only focused on the risk coming from one class of pathogen studied in high-biosafety research facilities, and did not account for the increase in the number of high-biosafety research facilities from 2017 to 2024. As of 2023, there were 51 BSL4 labs in operation, three under construction, and 15 planned, located across 27 countries.^12^ ^13^ Moreover, while the true number of BSL-3 labs worldwide is unknown, it is at least in the thousands, and likely increasing.^14^

Routine testing of lab workers for LAIs is not currently standard practice at high-biosafety research facilities. Discussions with current BSL-4 researchers revealed several potential explanations.^15^ First, lab managers may feel that routine testing is unnecessary given the existing safety precautions. Second, lab managers may be concerned that routine testing could be logistically burdensome and financially costly for individual labs. Third, lab managers may be concerned about the potential practical consequences of false positive results. Fourth, biosafety risk assessments performed by regulators, research funders, and lab managers do not always sufficiently focus their risk assessments on overall societal risk, rather than individual or lab-level occupational risk.^16^ Last, existing regulatory frameworks governing research with PPPs do not require or even discuss testing for accidental lab-acquired infection except in response to known exposures or symptoms.^17^

While routine testing of lab workers for LAIs is not currently standard practice at high-biosafety research facilities, similar activities have been undertaken before, indicating their potential feasibility. During the SARS-COV-2 pandemic, many workplaces were able to implement routine screening protocols of multiple tests per week, especially in healthcare settings and for high-revenue professional sports teams.^18^ Routine screening for community infections with SARS-COV-2 has also been performed in a high-biosafety research environment, Boston University’s combined BSL-4 and BSL-3 research center, the NEIDL,^19^ even as it also conducted gain-of-function experiments with the SARS-COV-2 virus.^15^

A key challenge with containing PPP LAIs is that for some pathogens, such as SARS-COV-2 and some strains of influenza, a substantial proportion of disease transmission is caused by asymptomatic or pre-symptomatic infections,^20,21^ which makes it difficult to contain an emerging outbreak solely with case-based interventions.^1^ Screening of individuals known to be at high risk of infection can help mitigate these challenges by identifying people with asymptomatic, pre-symptomatic, or mild symptomatic infections, which facilitates case-based interventions.^22,23^

This modeling study investigates the extent to which routine screening of lab workers in high-biosafety research environments for LAIs can reduce the risk of a catastrophic escape of a PPP similar to wild type SARS-COV-2 into the community. In particular, this analysis focuses on how the success of such a surveillance protocol would depend on test frequency, test sensitivity, and the average delay from a positive test result being taken until the effective isolation of the infected individuals.

## Methods

### Study Population

The target study population is the laboratory employees of a BSL-4 research facility and the surrounding community. To approximate this study population, we simulated a 1000 person contact network for a BSL-4 laboratory with 20 lab workers and 980 people in the surrounding community. A Barabasi-Albert network structure was chosen to allow a scale-free distribution of connections between individuals.^24^ The mean number of connections between individuals that were sufficiently close for transmission was set to eight per person, for both groups, but each individual’s number of connections was heterogenous due to the network structure, with a few individuals acting as network hubs.

### Intervention Strategies and Analytic Scenarios

Our goal was to explore the impact of a wide range of plausible test-and-isolate intervention strategies encompassing different values of test frequency, peak test sensitivity, and average delay from positive test to isolation. To achieve this, we conducted epidemic simulations encompassing a wide range of these parameter values using the contact network created for the study population. At the beginning of each simulation, we assumed a single randomly chosen lab worker developed a LAI. We then simulated disease progression, transmission, and other epidemiological outcomes for a subsequent 100 days, by which time the large majority of simulated epidemics reached the threshold for defining an outbreak or had otherwise ended.

Across simulations, the frequency of routine lab worker testing was systematically varied from zero tests per week up to seven tests per week in increments of one. Similarly, the value of peak test sensitivity was systematically varied from 50% to 100% in increments of 10%. Last, the average delay from positive test to isolation was systematically varied from zero days to 4 days in increments of one day. This produced 192 unique analytic scenarios.

### Data

Parameters characterizing the natural history of infection with wild-type SARS-CoV-2 were drawn from the clinical literature. Full definitions and values for the parameters are given in Table S1 in the Technical Appendix.

### Simulation model

We conducted this analysis with a discrete time stochastic network infectious disease model, with a one-day time-step. A schematic of the modelled health states and possible state transitions is given in Figure 2.1.

**Figure 2.1:**
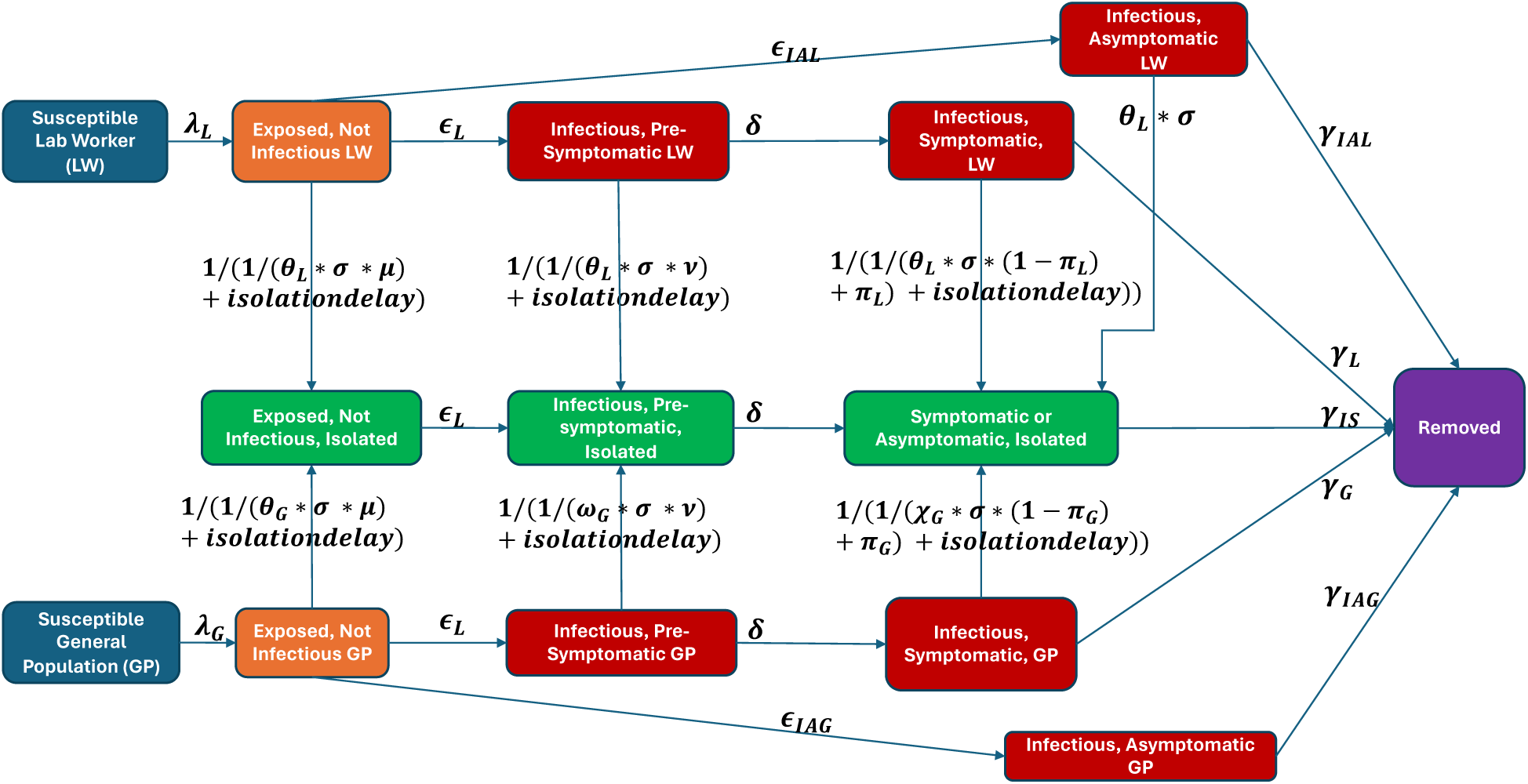
Model Structure

### Transmission Mechanisms

In this model, we assume that an infection is transmitted across a given susceptible-infectious edge with probability τ per time step *t*. In the simplest version of such a chained binomial model, the probability of any particular susceptible becoming infected in a given time-step is *p* = 1 − (1 − τ)*^y^*, where *y* is the number of infected contacts.^24^ The spread of epidemics on networks is influenced by both the mean number of contacts (edges) k, as well as the heterogeneity in that number, which is quantified by k’s relationship with *k*^2^, as evidenced by the equation for a simple chained binomial model, 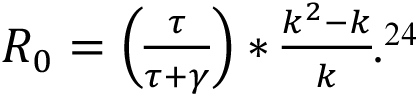 To enhance the model, the equation was modified to account for differential transmission by individuals with pre-symptomatic and asymptomatic infection relative to symptomatic disease, as well as the greatly reduced transmission of isolated infected contacts. This was done in several ways. First, the model includes a transmission offset **ζ** that is multiplied by the number of asymptomatic infected contacts to account for the diminished infectiousness of individuals with asymptomatic infection relative to individuals with symptomatic infection. Second, the model features a transmission offset ***ψ*** that is multiplied by the number of pre-symptomatic infected contacts to account for the diminished infectiousness of pre-symptomatic infectees relative to individuals with symptomatic infection. Third, the model includes a transmission offset ***i*** multiplied by the number of isolated infected contacts which represents the (small) fraction of transmission that isolated infectious individuals exhibit even while isolated, relative to non-isolated infectious individuals. The modified transmission equation is given in Table S1 in the Technical Appendix.

### Screening and Isolation

For every time step, lab workers were assumed to be tested with probability *θ_L_*. In cases when the tested individual was infectious and symptomatic, the person would be isolated with probability σ, the peak test sensitivity. If the tested individual was infectious and pre-symptomatic, the person would be isolated with probability σ ∗ *v*, with *v* representing the reduction in test sensitivity during the pre-symptomatic period. If the tested individual was infected but not yet infectious or even symptomatic, the person would be isolated with probability σ ∗ μ, with μ representing the heavily reduced test sensitivity during this period. In addition to test-induced isolation, the model incorporated baseline rates by which symptomatic lab workers and symptomatic general population members would isolate themselves even without testing positive, **π**_**L**_ and **π_G_** respectively. The model was programmed so that in all simulations, after the first positive test result was recorded, the testing cadence for lab workers increased to daily. Similarly, in all simulations after more than ten positive test results were recorded infected, symptomatic members of the general population became subject to a testing cadence of one test per week. To minimize model complexity, tests were assumed to be 100% specific, with no false positives.

### Study Outcome

The key outcome in this study was whether an outbreak of 50 or more infections occurs within 100 days of the initial infection. In sensitivity analyses we also explored whether 20 or more and 10 or more infections occurred within 100 days.

### Simulations and Statistical Analysis

We conducted one thousand simulations for each unique analytic scenario, for a total of 192,000 simulations. To execute in parallel the large number of simulations required, simulations were executed using the Harvard FAS Research Computing Cluster. As the simulations were conducted, summary data from each simulation were extracted and aggregated into a comprehensive analytic data frame. Once the comprehensive analytic data frame was created, a series of logistic regression analyses were performed to summarize the relationships between the probability of an outbreak of 50 or more and the test frequency, test sensitivity, and average isolation delay. The use of logistic regression to summarize these relationships was important because the magnitude of each relationship was unknown prior to running the simulations due to the inherent stochasticity and complex network structure of the model. Two versions of logistic regression analysis were performed: one without interaction terms between predictors (see Table 2.1) and one with interaction terms between predictors (see Table 2.2). The contact network simulation, epidemic simulations, and the statistical analysis were all conducted in the R programming language.

**Table 2.1:** Probability of an Outbreak of 50 or More Infections Under Various Testing Scenarios.

|  |  |  |
| --- | --- | --- |
| **Testing Scenario** | **Probability of Outbreak of 50+** | **Risk Ratio** |
| **No Routine Testing:** 0 tests per week | 0.42 | 1 |
| **Low-Intensity, Pessimistic:** 1 test per week, peak sensitivity of 50%, isolation delay of 2 days | 0.3 | 0.714 |
| **Low-Intensity, Optimistic:** 1 test per week, peak sensitivity of 80%, isolation delay of 1 day | 0.2 | 0.48 |
| **Moderate-Intensity, Pessimistic:** 2 tests per week, peak sensitivity of 50%, isolation delay of 2 days | 0.24 | 0.57 |
| **Moderate-Intensity, Optimistic:** 2 tests per week, peak sensitivity of 80%, isolation delay of 1 day | 0.16 | 0.38 |
| **High-Intensity, Pessimistic:** 5 tests per week, peak sensitivity of 50%, isolation delay of 2 days | 0.19 | 0.45 |
| **High-Intensity, Optimistic:** 5 tests per week, peak sensitivity of 80%, isolation delay of 1 day | 0.12 | 0.29 |

**Table 2.2:** Simple Logistic Model of Outbreaks of 50 or More Infections.

| **Table 2.2: Simple Logistic Model of Outbreaks of 50 or More Infections** |  |  |  |  |  |
| --- | --- | --- | --- | --- | --- |
|  | Odds Ratios | Lower Bound 95% CI | Upper Bound 95% CI | Z-Score | P-Value |
| Frequency | 0.800 | 0.796 | 0.805 | -82.124 | <0.001 |
| Sensitivity | 0.990 | 0.989 | 0.990 | -29.97 | <0.001 |
| Isolation Delay | 1.155 | 1.143 | 1.168 | 27.050 | <0.001 |
| Intercept | 0.377 | 0.367 | 0.386 | -76.291 | <0.001 |

### Sensitivity Analyses

To ensure the analysis results were robust across different thresholds for defining whether an outbreak had occurred, three thresholds were selected for analysis: outbreaks with 50 or more infections, 20 or more infections, and 10 or more infections. In addition, among the subsets of epidemics that exceeded these thresholds, we calculated the proportion of epidemics that were still growing, as indicated by a positive exponential growth rate over the final 20 days. To ensure the pathogen behaved as expected given the epidemiological parameters meant to approximate wild-type SARS-COV-2, we calculated the observed basic reproductive number R0 in the second week after the initial infection in simulations when there was no routine testing, and confirmed that it was consistent with an R0 of 2.0, similar to the WHO’s R0 estimate of 1.95 for the very early stages of the SARS-COV-2 pandemic in China in January 2020,^25^ as well as to the 2.28 R0 value estimated from the early part of the SARS-COV-2 outbreak on the Diamond Princess passenger cruise ship in 2020.^26^

### Human subjects considerations

No human subjects were involved in this research.

## Results

### Risk Reductions Under Plausible Testing Scenarios

By simulating a range of plausible scenarios, we found that a screening intervention with a range of test frequencies, sensitivity, and average delay from positive test to isolation could substantially reduce the probability of an outbreak of 50 or more infections (see Table 2.1 and Figure 2.2). The probability of such an outbreak in the simulations of the baseline scenario with no routine testing was 0.42. In simulations capturing a low-intensity, pessimistic testing scenario of 1 test per week, peak sensitivity of 50%, and average isolation delay of 2 days, the probability of a 50+ infection outbreak fell by 29% (RR: 0.71) to 0.30, compared with the baseline risk of 0.42. In simulations capturing a low-intensity, optimistic testing scenario of 1 test per week, peak sensitivity of 80%, and average isolation delay of 1 day, the probability of a 50+ infection outbreak fell by 52% (RR: 0.48), to 0.2, compared with the baseline risk of 0.42. In simulations capturing a moderate-intensity, pessimistic scenario of 2 tests per week, peak sensitivity of 50%, and average isolation delay of 2 days, the probability of an outbreak fell by 43% (RR: 0.57), to 0.24, compared with the baseline risk of 0.42. In simulations capturing a moderate-intensity, optimistic scenario of 2 tests per week, peak sensitivity of 80%, and average isolation delay of 1 day, the probability of an outbreak fell by 62% (RR: 0.38), to 0.16, compared with the baseline risk of 0.42. In simulations capturing a high-intensity, pessimistic scenario of 5 tests per week, peak sensitivity of 50%, and average isolation delay of 2 days, the probability of an outbreak fell 55% (RR: 0.45), to 0.19, compared with the baseline risk of 0.42. Last, in simulations capturing a high-intensity, optimistic scenario of 5 tests per week, peak sensitivity of 80%, and average isolation delay of 1 day, the probability of an outbreak fell by 71% (RR: 0.29), to 0.12, compared with the baseline risk of 0.42.

**Figure 2.2:**
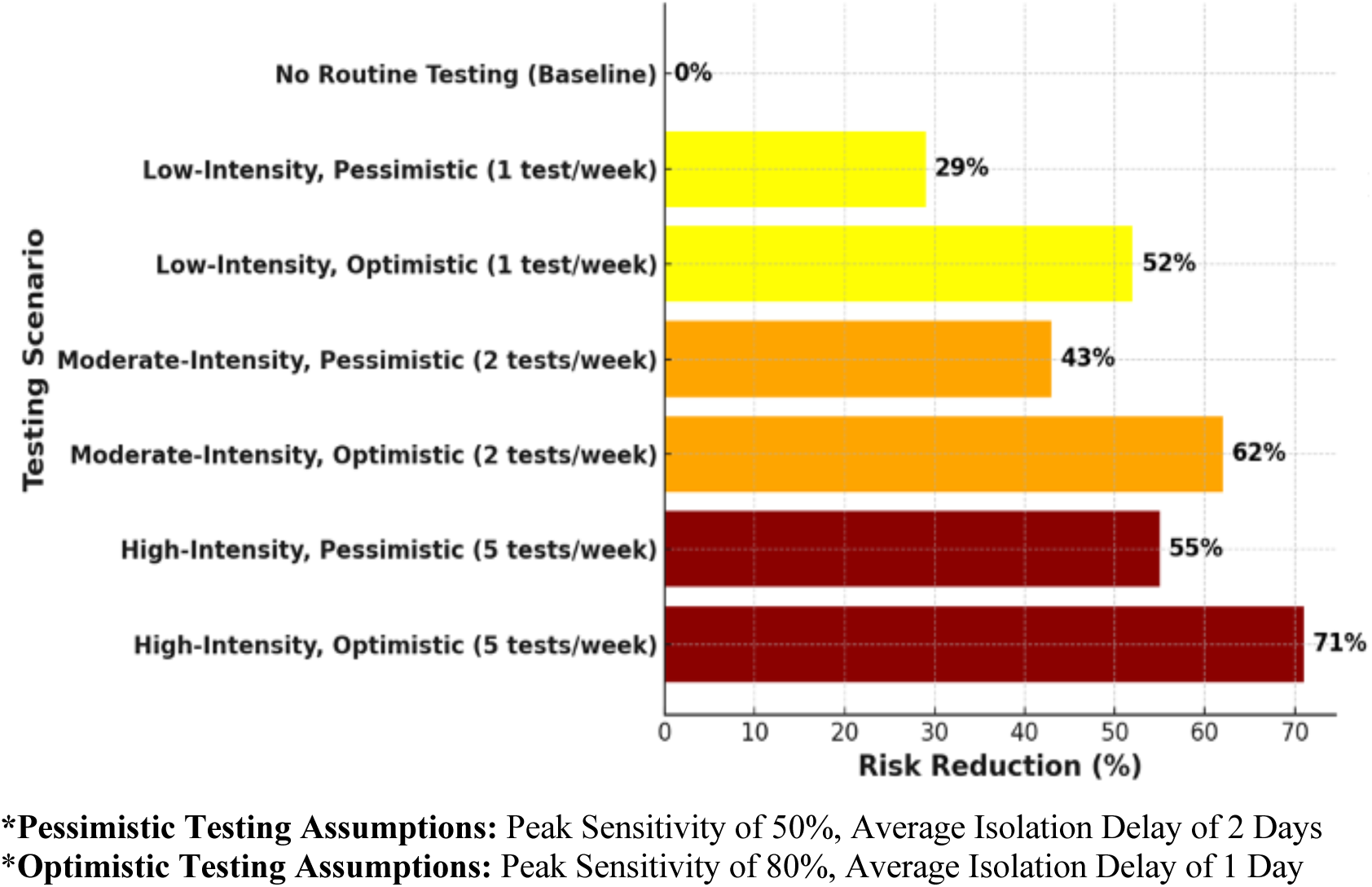
**Risk Reduction Under Various Testing Scenarios\***

### Factors Predicting Outbreaks

At the outbreak threshold of 50 or more infections, each of the three predictors was found to have a significant independent effect on the likelihood of an outbreak occurring, in both the models with and without interaction terms, but the strongest effect was observed for test frequency. In the simple logistic model, each additional day per week of test frequency was associated with a 20% reduction in the odds of an outbreak (see Table 2.2 and Figure 2.3). In the model with interaction terms, the effect of increased test frequency was moderated by an interaction effect with the average isolation delay (see Table 2.3). In the logistic model with interactions (see Table 2.2), increasing test frequency to one, two, and five days per week relative to a baseline of no routine testing when peak test sensitivity was 75% (the mean value in the simulations) and there was no isolation delay reduced the odds of an outbreak by 24%, 43%, and 75%, respectively. As the average isolation delay increased, the increase in test frequency led to smaller reductions in the odds of an outbreak. For example, in a scenario with a relatively long average isolation delay of two days and a mean test sensitivity of 75%, increasing test frequency to one, two, and five days per week relative to a baseline of no testing reduced the odds of an outbreak by 19%, 35%, and 66%, respectively (see Table 2.2). In the model with interaction terms, the effect of increased test frequency was modestly magnified by an interaction effect with test sensitivity, such that the effect of increased testing was greater at higher levels of test sensitivity.

**Figure 2.3:**
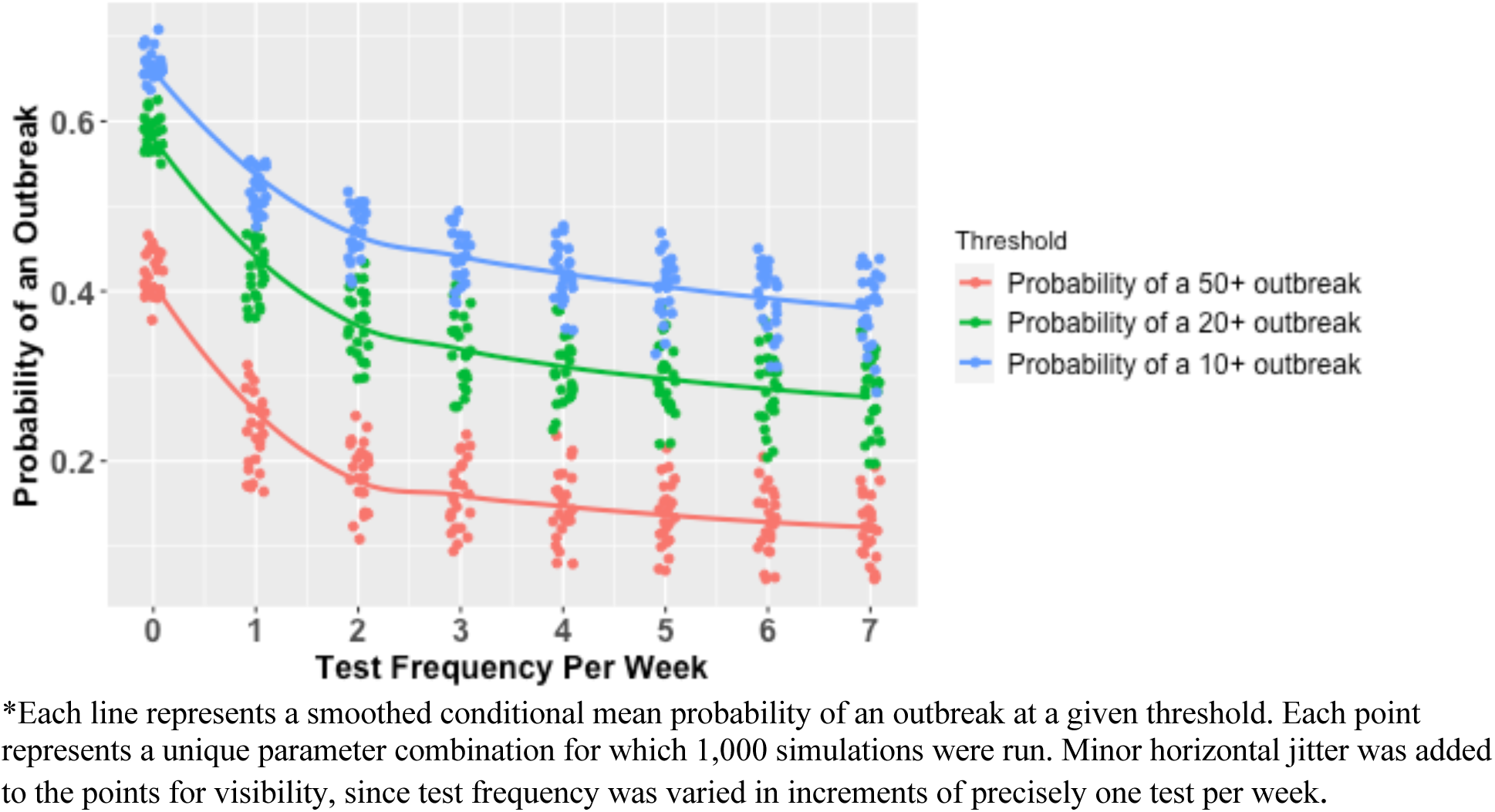
Marginal Probability of an Outbreak of Various Sizes by Test Frequency*

**Table 2.3:** Logistic Model with Interactions of Outbreaks of 50 or More Infections.

|  |  |  |  |  |  |
| --- | --- | --- | --- | --- | --- |
|  | Odds Ratios | Lower Bound 95% CI | Upper Bound 95% CI | Z-Score | P-Value |
| Frequency | 0.757 | 0.749 | 0.764 | -56.963 | <0.001 |
| Sensitivity | 0.992 | 0.991 | 0.994 | -10.528 | <0.001 |
| Isolation Delay | 1.057 | 1.040 | 1.075 | 6.450 | <0.001 |
| Frequency X Sensitivity | 0.998 | 0.998 | 0.999 | -9.612 | <0.001 |
| Frequency X Isolation Delay | 1.033 | 1.028 | 1.038 | 13.281 | <0.001 |
| Sensitivity X Isolation Delay | 1.001 | 1.000 | 1.002 | 3.155 | <0.001 |
| Intercept | 0.437 | 0.423 | 0.451 | -49.940 | <0.001 |

In the simple logistic model, a 10-percentage point increase in test sensitivity was associated with a 10% reduction in the odds of an outbreak (see Table 2.2 and Figure 2.4). In the model with interaction terms, the effect of increased test sensitivity was slightly moderated by an interaction effect with the average isolation delay (see Table 2.3). As the average isolation delay increased, the increase in test sensitivity led to smaller reductions in the odds of an outbreak.

**Figure 2.4:**
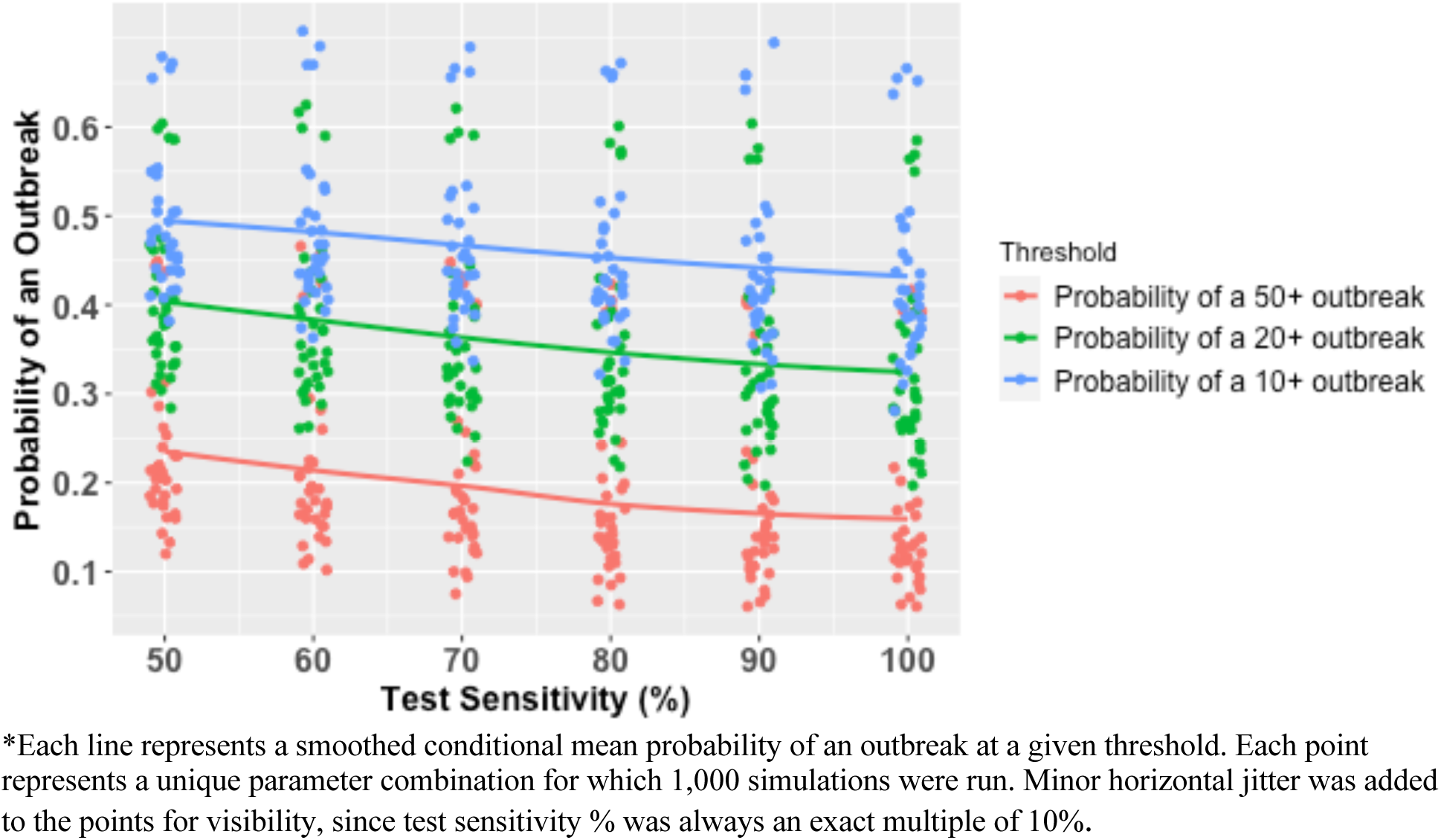
Marginal Probability of an Outbreak of Various Sizes by Test Sensitivity

Similarly, the effect of increased test sensitivity was somewhat magnified by an interaction effect with test frequency, such that the effect of increased test sensitivity was greater at higher levels of test frequency.

In the simple logistic regression model, an increase of one day in the average delay from a positive test to effective isolation was associated with a 15.5% increase in the odds of an outbreak (see Table 2.2 and Figure 2.5). In the model with interaction terms, isolation delays influenced the likelihood of outbreaks partially through an attenuated main effect (OR of 1.057 compared to 1.155) and partially through interactions with test frequency and test sensitivity that attenuated the protective effects of increasing those variables when isolation delays were longer (see Table 2.3, Table 2.4, and Table 2.5).

**Figure 2.5:**
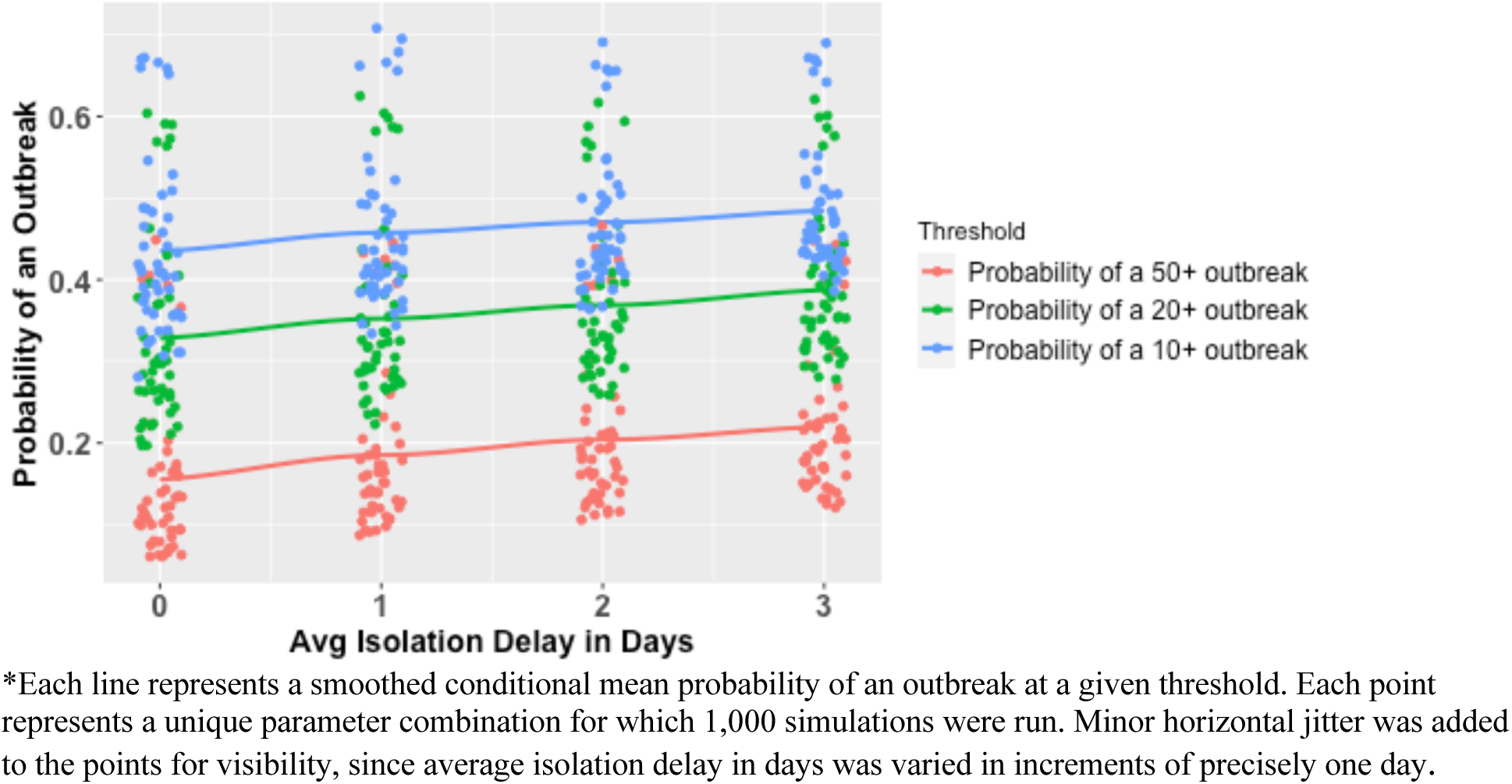
Marginal Probability of an Outbreak of Various Sizes by Average Isolation Delay

**Table 2.4:** Odds Ratios for Test Frequency by Average Isolation Delay (From Logistic Model with Interactions for Outbreaks of 50 or More Infections)

|  |  |  |  |  |  |  |  |  |
| --- | --- | --- | --- | --- | --- | --- | --- | --- |
| Tests Per Week | 0 | 1 | 2 | 3 | 4 | 5 | 6 | 7 |
| OR with Avg Isolation Delay of 0 Days, with 95% CI | 1.00 | 0.757<br>(0.749 to 0.764) | 0.573<br>(0.561 to 0.584) | 0.434<br>(0.420 to 0.446) | 0.328<br>(0.315 to 0.341) | 0.249<br>(0.236 to 0.260) | 0.188<br>(0.177 to 0.199) | 0.142<br>(0.132 to 0.152) |
| OR with Avg Isolation Delay of 1 Day, with 95% CI | 1.00 | 0.782<br>(0.770 to 0.793) | 0.611<br>(0.593 to 0.629) | 0.478<br>(0.456 to 0.499) | 0.374<br>(0.351 to 0.396) | 0.292<br>(0.271 to 0.314) | 0.229<br>(0.208 to 0.249) | 0.179<br>(0.16 to 0.197) |
| OR with Avg Isolation Delay of 2 Days, with 95% CI | 1.00 | 0.808<br>(0.792 to 0.823) | 0.653<br>(0.627 to 0.678) | 0.527<br>(0.496 to 0.558) | 0.426<br>(0.393 to 0.459) | 0.344<br>(0.311 to 0.378) | 0.278<br>(0.246 to 0.311) | 0.224<br>(0.195 to 0.256) |
| OR with Avg Isolation Delay of 3 Days, with 95% CI | 1.00 | 0.834<br>(0.814 to 0.854) | 0.696<br>(0.662 to 0.730) | 0.581<br>(0.539 to 0.624) | 0.485<br>(0.438 to 0.533) | 0.405<br>(0.357 to 0.455) | 0.338<br>(0.29 to 0.389) | 0.282<br>(0.236 to 0.333) |

**Table 2.5:**
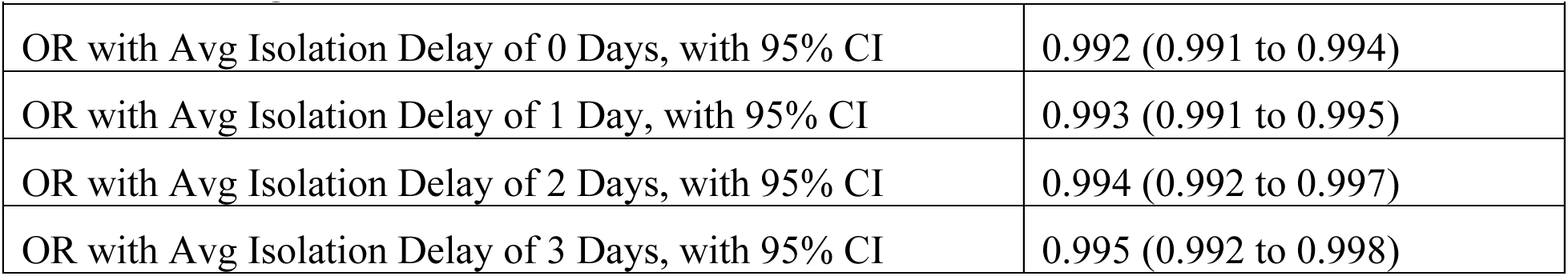
Odds Ratios for a 1 Percentage Point Increase in Test Sensitivity by Average Isolation Delay (From Logistic Model with Interactions for Outbreaks of 50 or More Infections)

These results were qualitatively similar at the three outbreak thresholds of 50, 20 and 10 or more infections, as well as across models both without and with interaction terms. Among the subsets of simulations with 50 or more cumulative infections, 20 or more cumulative infections, and 10 or more cumulative infections, respectively, the percentages of epidemics that were still growing as indicated by a positive exponential growth rate over the final 20 days were 32%, 36%, and 32% (See Table 2.6).

**Table 2.6:** Proportion of Outbreaks Of Various Thresholds Still Growing at 100 Days.

|  |  |  |
| --- | --- | --- |
| Proportion of outbreaks of 50+ growing at 100 days | Proportion of outbreaks of 20+ growing at 100 days | Proportion of outbreaks of 10+ growing at 100 days |
| 0.32 | 0.36 | 0.32 |

## Discussion

In this study, we found that the risk reduction from routine screening for LAIs with PPPs be substantial under a wide variety of assumptions about test frequency, test sensitivity, and the average delay from positive test to effective isolation, as well as at various thresholds for defining an outbreak. For example, under our logistic model with interaction (Table 2.2), assuming peak test sensitivity of 75% and average isolation delays of between zero to two days, increasing test frequency to one, two, and five days per week relative to a baseline of no testing reduced the odds of an outbreak of 50 or more by 19-24%-, 35-43%, and 66-65%, respectively.

It is worth noting that the baseline risk of an outbreak of 50 or more infections conditional on the first lab worker’s infection in the absence of routine testing was high, at 0.42, and while routine testing and isolation interventions substantially reduced the risk, it still remained significant. For example, even under a high-intensity, optimistic scenario of 5 tests per week, peak sensitivity of 80%, and average isolation delay of 1 day, where the probability of an outbreak was reduced by 71% (RR: 0.29) compared with the baseline risk of 0.42 to a new risk of 0.12, the risk remained far from 0. Among the subsets of simulations with 50 or more cumulative infections, 32% of were still growing over the final 20 days and risked becoming a serious epidemic. This suggests the difficulty of containing the epidemic with the modeled interventions once sustained community transmission is underway.

Test frequency was found to be the most important factor in influencing outbreak risk. This is notable because test frequency is a highly modifiable feature of a screening program’s design. Testing just one or two days per week yielded substantial risk reductions in this modeling study, with even greater cumulative risk reductions achievable at higher testing frequencies. While increased test sensitivity was also found to influence outbreak risk, this effect is of modest practical importance for two reasons. First, the incremental reduction in the odds of an outbreak resulting from a given increase in test sensitivity was moderate. Second, in practice, test sensitivity can only be improved within a limited range. The length of average delay from a positive test to effective isolation was observed to be important because longer isolation delays are associated with higher outbreak risks. A longer delay from a positive test to effective isolation will reduce the protective value of greater test frequency and sensitivity by reducing the proportion of an infected person’s infectious time that they spend in isolation.

There may be tradeoffs in program design between optimizing test frequency, test sensitivity, and isolation delays. It would be unwise to infer generalized rules from one modeling study about how all potential tradeoffs in routine screening program design between different test characteristics should be navigated. However, this modeling study does offer evidence that when faced with such tradeoffs, program designers will most likely achieve the greatest benefits by prioritizing high test frequency, within reason.

No new technology would likely be necessary to conduct routine testing for LAIs in high-biosafety research environments. Sensitive, commercially available molecular PCR tests already exist for the major PPPs that are studied in high-biosafety research environments that are of greatest concern, particularly coronaviruses and influenzas.^27^ ^28^ Moreover, the key regions of pathogens’ genetic material that are necessary to trigger a positive test result are generally conserved even for pathogens modified in the laboratory.^27^ ^28^ If individual research facilities sought more specific tests that could distinguish between viral strains modified in a lab versus those circulating in nature, they would likely have the in-house expertise to develop them.^27^ ^28^ To minimize costs and logistical burdens, these more specific tests could be held in reserve for follow-up investigation after an initial positive test result from a commercially available test.

This study has several limitations. First, it does not quantify the likely additional risk reductions that could be achieved by layering additional interventions on top of routine testing and isolation of infected lab workers, such as contact tracing and the use of medical countermeasures. Second, while this study can inform us regarding the relative reductions in the risk of escape of a PPP into the community (conditional on an accidental infection of a lab worker) that could be achieved through a routine testing intervention, this study cannot tell us the absolute risk reductions such an intervention would achieve, because the baseline absolute risks of such accidental infections have not been well-established by the literature. Third, this study does not demonstrate the cost-effectiveness of the interventions studied. Fourth, this study only assessed the efficacy of the intervention for a PPP with characteristics similar to wild-type SARS-COV-2.

Additional research will be needed to answer related questions. First, future research should explore how the effectiveness of such a routine screening system might be influenced by pathogens’ epidemiologic characteristics. Second, future research should attempt to quantify the financial and human resource costs of implementing such a screening program, as well as its cost-effectiveness. Third, future research should address other policy questions such as who would finance and operate such a program and what regulatory framework it would operate under. Fourth, future research should address likely implementation questions that would arise such as how to optimally design a screening system under resource constraints and how to maximize fidelity in its implementation. Last, future research should work to more firmly establish the baseline level of risk posed by laboratory escapes of PPPs, perhaps by using alternative approaches like anonymous serology that can measure undetected past exposures.

The large potential benefits of routine screening identified in this modeling study provide evidence in favor of both further modeling research as well as implementation research in the context of a pilot routine screening program. As the evidence base develops, policymakers may wish to consider updating biosafety regulations or guidelines to promote routine screening for LAIs for facilities working with PPPs.

## Conclusions

This modeling study finds that routine screening of lab workers in high-biosafety research environments for accidental lab-acquired infections (LAIs) coupled with isolation of infected workers could substantially reduce the risk of a catastrophic escape of a potential pandemic pathogen (PPP) similar to wild type SARS-COV-2, with the extent of risk reduction dependent on the intensity of the testing regime. Test frequency was found to be the most important factor influencing outbreak risk, while test turnaround time and sensitivity also influenced outbreak risk. The risk reduction from routine screening remained large under a range of assumptions about test frequency, test sensitivity, and the average delay from positive test to effective isolation, as well as at various thresholds for defining an outbreak.

## Data Availability

All data produced in the present study are available upon reasonable request to the authors.

